# No evidence of viral polymorphisms associated with Paediatric Inflammatory Multisystem Syndrome Temporally Associated With SARS-CoV-2 (PIMS-TS)

**DOI:** 10.1101/2020.07.07.20148213

**Authors:** Juanita Pang, Florencia A.T. Boshier, Nele Alders, Garth Dixon, Judith Breuer

## Abstract

Generally, children and teenagers do not become seriously ill with COVID-19. However, in countries with high rates of coronavirus disease, children with the syndrome COVID-19 associated inflammation syndrome referred to as PIMS-TS have been reported. Similarities noted between SARS-CoV-2 Spike protein sequences and those of other super antigens has prompted the suggestion that this might be the mechanism by SARS-CoV-ST triggers PIMS-TS. It has also been suggested that the D614G variant found more commonly in the US and across European countries may explain why PIMS-TS appears to be common in these countries. Here we analysed viral sequences from 13 paediatric COVID-19 patients of whom five were diagnosed with PIMS-TS. This is the first characterisation of viruses from PIMS-TS patients. In contrast to what has been hypothesised, we found no evidence of unique sequences associated with the viruses from PIMS-TS patients.

---

Severe illness and death due to SARS-CoV-2 infection in children is rare. However, a small number of cases of shock and multisystem inflammation have been reported in children who have either been tested positive for SARS-CoV-2 (by PCR or serology) or had epidemiological links to it. This new syndrome is called the paediatric inflammatory multisystem syndrome temporally associated with SARS-CoV-2 (PIMS-TS)^1^.

The exact pathogenesis of PIMS-TS is as yet unknown. However, it has been suggested that part of the SARS-CoV-2 viral spike (S) protein may resemble a superantigen which could drive the development of PIMS-TS and triggers a cytokine storm in adults^2^. Specifically, polymorphic residues in S including A831V and D839Y/N/E^2^ which are predicted to enhance binding affinity to the TCR have been observed in lineages circulating in Europe and North America, where most PIMS-TS cases have been described. In addition, the 614G Spike protein polymorphism may be associated with increased transmission and altered SARS-CoV-biology^3^.

To examine whether viral sequence variation might contribute to the pathogenesis of PIMS-TS, we sequenced SARS-CoV-2 from children hospitalised for COVID-19 in London between late-March and mid-May 2020. Of 61 hospitalised children with COVID-19, 36 were diagnosed with PIMS-TS, 11 of whom were positive for SARS-CoV-2 viral RNA. Full length SARS-CoV-2 genome sequences were obtained from 5 PIMS-TS children and 8 non-PIMS-ST children using SureSelect^XT^ target enrichment and Illumina sequencing. Reads generated were quality checked and mapped to the SARS-COV-2 reference genome (NC_045512) from GenBank. Sequences are available on GISAID (Accession ID: EPI_ISL_479777 to EPI_ISL_479789).

We constructed a maximum likelihood phylogeny of these sequences and 130 SARS-CoV-2 sequences generated from community cases across North London (Figure A). There was no clustering of viral sequences from PIMS-TS patients or non PIMS-TS patients in relation to other local sequences.

Using the reference genome (NC_045512), we observed no single nucleotide polymorphisms (SNPs) unique to the PIMS-TS or to the other childhood cases and no difference in the distribution of SNPs between PIMS-TS, non PIMS-TS and community cases (Figure B). All childhood cases were D839 and A831 as were all of the locally circulating samples. The majority of PIMS-ST (3/5), non PIMS-ST (6/8) and community cases (118/130) were 614G positive.

Overall, the data suggest that the viruses causing PIMS-TS in our patients are representative of locally circulating SARS-CoV-2. We found no evidence for an association of PIMS-TS with the presence of new or unusual sequence polymorphisms.

**Figure:**
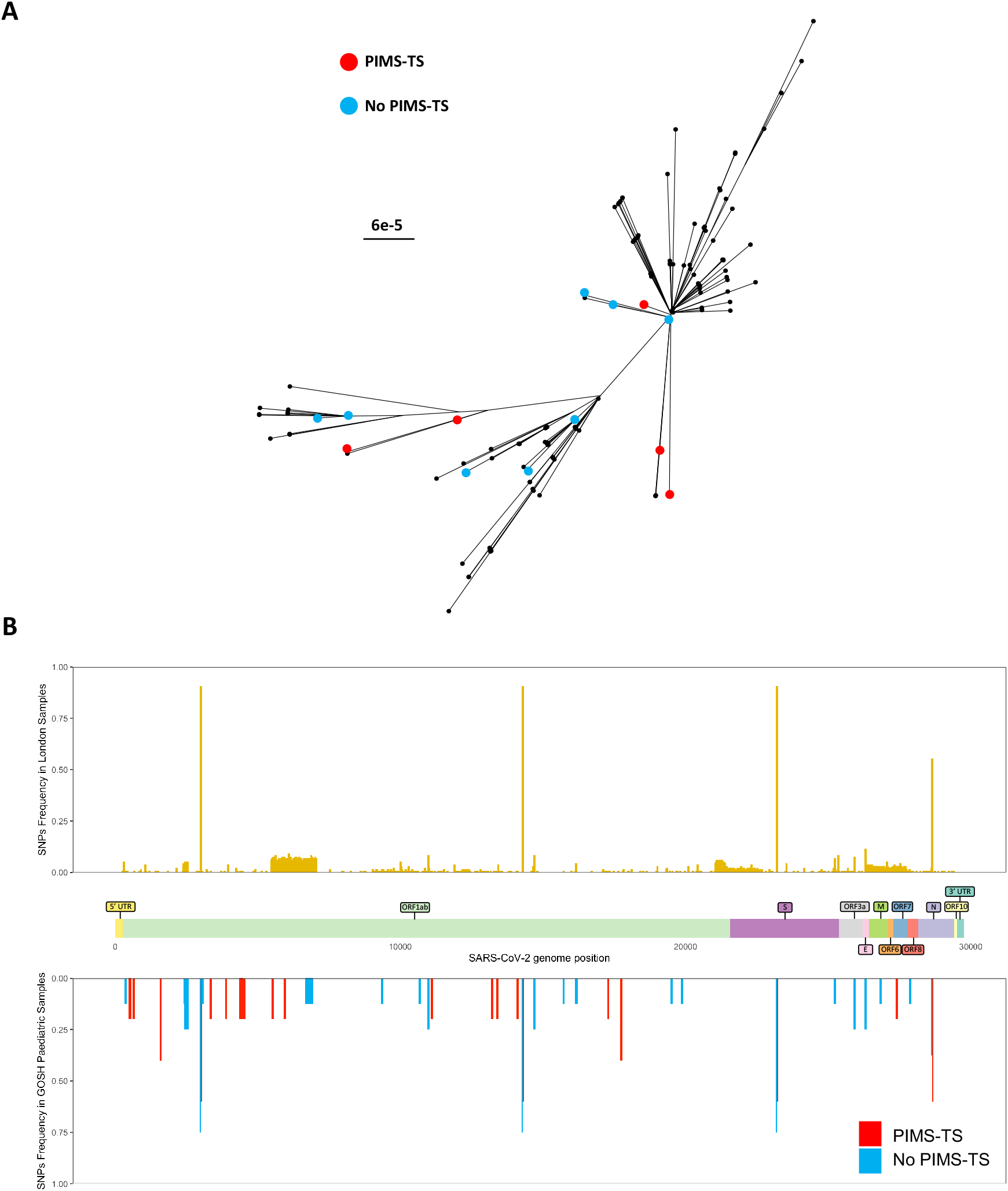
Characteristics of SARS-CoV-2 sequences from children with and without PIMS-TS. (A) Phylogenetic tree of sequences analysed in this study. Tips coloured in red = from PIMS-TS children, blue = from non-PIMS-TS children, black = other sequences from London with no association to PIMS-TS. (B) Frequency of occurrence of single nucleotide polymorphisms. Top: 130 London Samples in yellow. Bottom: 13 Paediatric samples from GOSH, red = 5 PIMS-TS samples, blue = 8 non PIMS-TS samples. The x-axis is annotated with a map of the reading frames in the viral genome.

## Data Availability

all the sequences have been submitted to COG-UK and are freely available in the MRC CLIMB database

## Acknowledgements

This work was supported by COG-UK, The James Black Charitable Foundation and the UCLH and GOSH NIHR biomedical research centres. JP is supported by the Rosetrees Foundation and FTB by a Wellcome Trust Collaborative Award to JB. We thank Richard Goldstein, Kathryn Harris, Julianne Brown, Jack Lee, Rachel Williams, Helena Tutill and Sunando Roy for their contribution to this work. We should also like to acknowledge the contribution of the UCL Pathogen Genomics Unit, UCL Genomics, and the Great Ormond Street Hospital Departments of Infectious Disease, and Microbiology, Virology and Infection Control.

We declare no competing interests.

## Notes

### Competing Interest Statement

The authors have declared no competing interest.

### Funding Statement

this work was supported by the John Black Charitable foundation

### Author Declarations

The National Research Ethics Service (NRES) Committee London Fulham (REC reference: 17/LO/1530)

